# The Applications of Laser Interstitial Thermal Therapy and Machine Learning in Neurosurgery: A Systematic Review

**DOI:** 10.1101/2023.12.21.23300384

**Authors:** Andrew Bouras, Dhruv Patel, Nitin Chetla

## Abstract

**Background:** The incorporation of Machine Learning (ML) into Laser Interstitial Thermal Therapy (LITT) represents a significant advancement in minimally invasive neurosurgery, particularly for treating brain tumors, vascular malformations, and epileptogenic foci. This systematic review focuses on evaluating the integration and impact of ML in enhancing the efficacy, precision, and outcomes of LITT in neurosurgical procedures.

**Methods:** An exhaustive search was conducted in major scientific databases for studies from 2015 to 2023 that specifically focused on the application of ML in LITT. The review assessed the development and implementation of ML algorithms in surgical planning, outcome prediction, and postoperative evaluation in LITT. Rigorous inclusion criteria were applied to select studies, and a combination of meta-analysis and qualitative synthesis was used to analyze the data.

**Results:** The review synthesizes findings from a range of studies, including retrospective analyses and initial clinical trials. It highlights the role of ML in enhancing the selection criteria for LITT, optimizing surgical approaches, and improving patient-specific outcome predictions. While LITT showed favorable results in treating non-resectable lesions, the integration of ML was found to potentially refine these outcomes further. However, challenges such as the need for larger sample sizes, standardization of ML algorithms, and validation of these methods in clinical settings were noted.

**Conclusions:** The integration of ML into LITT procedures marks a promising frontier in neurosurgery, offering potential improvements in surgical accuracy and patient outcomes. The evidence suggests a need for continued development and rigorous testing of ML applications in LITT. Future research should focus on the refinement and validation of ML algorithms for wider clinical adoption, ensuring that technological advancements align with patient safety and treatment efficacy.

## 1 Introduction

### 1.0.1 Background on LITT and machine learning in neurosurgery

The advent of minimally invasive techniques has driven the investigation and adoption of laser interstitial thermal therapy (LITT) for a range of neurosurgical indications, namely deep-seated, surgically inaccessible, tumor resection and medically refractory epilepsy (Melnick et at., 2021) (Zeller et al., 2021). In addition to laser ablation, LITT utilizes laser energy delivered via an inserted probe to induce thermal destruction of pathological tissue (Salem et al., 2019). By allowing ablation of challenging lesions with reduced morbidity compared to open surgery, LITT enhances the treatment options for brain tumors as well as epileptogenic, vascular, and other intracranial pathologies. In parallel, the integration of machine learning (ML) methods for surgical planning, predictive modeling, and monitoring of outcomes is transforming techniques to optimize and assess LITT implementation.

LITT provides targeted treatment of lesions in eloquent or deep brain areas which previously had high risk for surgical morbidity. The well-circumscribed ablation zones and real-time monitoring of LITT allow for the controlled destruction of diseased tissue while protecting the healthy surrounding brain (Thomas et al., 2016). Such properties have resulted in the growth of LITT across practices. ML analytics leveraging advanced algorithms on diverse patient data have shown early success in automating complex planning tasks, forecasting LITT outcomes, and tracking effectiveness across sites (Li et al., 2019; Kolasa et al., 2023). However, variability in techniques, patient selection, and study designs have led to calls for further research and guidance on best practices for LITT adoption and ML integration (Viozzi et al., 2021; Miranda de Souza et al., 2023).

### 1.0.2 Rationale/objectives for systematic review

Given the rapid evolution of LITT implementation and ML integration in neurosurgery, a synthesis of the evidence base is needed to inform appropriate adoption and future research. However, variability across study protocols, patient cohorts, applications, and measures of efficacy and safety poses challenges in creating unified guidelines. Therefore, this review aimed to systematically gather the literature related to the use of LITT and ML analytics in neurosurgical settings. The goals of this systematic examination were multi-fold, seeking to synthesize the extent of evidence regarding several aspects crucial to these emerging modalities. For LITT specifically, the review consolidates findings on procedural guidance, clinical effectiveness in varied intracranial pathologies, and the known safety and complication rates that influence its risk/benefit profile. Furthermore, the review summarizes the implementations of ML techniques across the LITT treatment workflow from preoperative planning and predictive algorithms to methods for monitoring outcomes. Finally, scrutinizing the state of literature allows the identification of gaps in adoption practices, variability in evidence quality, and areas requiring future research efforts or consensus guidelines, both for clinical translation as well as optimal technological integration for LITT and ML tools. By evaluating these diverse facets, this review offers insights into the use of these technologies in neurosurgery.

## 2 Methods

### 2.1 Search strategy

An exploration of scientific databases was conducted, notably PubMed, IEEE Xplore, Google Scholar, and ScienceDirect, to encompass a wide range of literature pertinent to the applications of LITT and integration of machine learning in neurosurgery. Search terms such as “LITT,” “laser ablation,” “neurosurgery,” “machine learning,” “surgical planning,” and “predictive modeling” were used to capture relevant studies effectively. The search spanned the literature from January 2015 to December 2023 to target recent advances. Study selection began by screening titles and abstracts for relevance to LITT and ML techniques in neurosurgical contexts, followed by full-text review meetings aimed at clinical efficacy, safety profiling, and ML implementations related to LITT procedures. The focus was on incorporating high-quality primary research in peer-reviewed journals.

### 2.2 Eligibility criteria

Ineligible studies were excluded based on conference papers, reviews, non-primary research focus, or irrelevant topics of interest. Data extraction included noting study goals, methods, key findings, and conclusions, facilitating the qualitative synthesis of trends and themes related to LITT and ML adoption. Finally, the critical appraisal of individual studies’ limitations and biases ensured a comprehensive and impartial overview to inform future research directions. This methodology has provided insights into the evolving landscape of these technologies in neurosurgery.

### 2.3 Study selection and data extraction

The selected studies were evaluated for their objectives, methodologies, key findings, and overall conclusions. This allowed for a thorough understanding of each study’s contribution to the field. Information from these studies was compiled, with an emphasis on identifying overarching trends, patterns, and themes in the application and development of ML within LITT procedures.

### 2.4 Study Quality Assessment

Each study was critically appraised for methodological soundness, potential biases, and limitations. This step was crucial to ensure the integrity and reliability of the review. The assessment aimed to provide an objective overview of the current state of research, acknowledging areas of strength and weakness in the existing literature.

### 2.5 Data analyses

A qualitative synthesis approach was adopted to analyze the findings from the included studies. This method facilitated an in-depth understanding of the nuanced roles and impacts of ML in LITT. Special attention was given to how ML applications have influenced surgical outcomes, patient safety, and the overall efficacy of LITT procedures.

## 3 Results

### 3.1 Overview of studies on LITT

LITT has emerged as a transformative, minimally invasive technique for the treatment of various intracranial lesions, including brain tumors and other abnormalities (Johnson et al., 2022). This innovative approach uses laser energy delivered through a slender probe to induce thermal damage and coagulative necrosis in targeted lesions (Salem et al., 2019). Commonly referred to as laser ablation, LITT aims to optimize tumor control while reducing invasiveness and related morbidity, thereby presenting a shift in neurosurgical practices (Miranda de Souza et al., 2023).

LITT’s minimally invasive nature contrasts sharply with traditional open surgery, avoiding the need for craniotomy access. Studies such as that by Thomas et al. (2016) highlight its advantages, including shorter hospitalizations and faster recovery times. Furthermore, LITT’s compatibility with other treatments such as radiation or chemotherapy broadens its applicability in neuro-oncology.

The synthesis of the current evidence aims to evaluate the implementation of this technology in brain tumor and lesion ablation. Holste and Orringer provided a comprehensive review of the use of LITT for medically intractable lesions and reported favorable 3-year outcomes in terms of safety, efficacy, and extent of ablation in 252 patients with various pathologies (Holste and Orringer, 2019). Additionally, a systematic analysis by Damante et al. specifically focused on LITT utilization and outcomes for recurrent brain metastases, finding median progression-free survival rates of 7-8 months in over 300 cases across 10 studies (Damante et al., 2022).

The intersection of LITT with artificial intelligence (AI) and ML presents new prospects for enhancing treatment outcomes (Li et al., 2019). ML’s role extends across the LITT treatment spectrum and includes preoperative planning, intraoperative guidance, and postoperative assessment. Techniques such as random forests, support vector machines, and neural networks have been instrumental in optimizing treatment parameters and modeling expected outcomes (Kolasa et al., 2023).

For instance, ML algorithms have been pivotal in analyzing patient images and data, offering tailored recommendations for laser probe trajectories, or predicting ablation extents (Li et al., 2019). Another significant application is in prognostic assessments, where ML aids in evaluating the factors influencing progression-free survival post-LITT (Souza et al., 2023). These advancements underscore the potential of ML to refine LITT’s precision and overall efficacy of LITT in diverse clinical settings.

### 3.2 Mechanism, technique, imaging, histology

LITT operates on the principle of delivering focused laser light energy through an optical fiber, inducing hyperthermic ablation of the targeted brain tissue (Salem et al., 2019). This process, central to LITT’s therapeutic efficacy of LITT, involves elevating tissue temperatures to a range–42-60°C, resulting in irreversible cellular damage and coagulative necrosis.

The delivery of this laser energy is achieved using advanced LITT systems, which have evolved significantly over time. These systems facilitate the MRI-guided placement of a cooled laser probe into the lesion, ensuring precision and minimizing damage to adjacent healthy tissues. The cooling mechanism of the probe is critical because it allows for sustained release of therapeutic levels of heat without overheating the probe itself.

A pivotal aspect of LITT is the integration of simultaneous thermal MRI. This technology enables neurosurgeons to visualize the extent of tissue ablation in real time, which is a significant advancement over the traditional methods (Miranda de Souza et al., 2023). Real-time imaging not only enhances the precision of the procedure but also significantly increases its safety profile by preventing unintended damage to adjacent brain structures.

Characteristic post-LITT imaging findings include a central zone of necrosis surrounded by a variable degree of edema, which typically resolves over time. Histologically, the ablated zones are marked by complete necrosis with sharply demarcated borders between the dead tissue and viable surrounding brain tissue (Thomas et al., 2016). These distinct histological features are pivotal for understanding LITT’s efficacy of LITT in targeting tumor cells while preserving the surrounding brain structures.

Understanding the mechanism of LITT, combined with advancements in techniques and imaging, is crucial for neurosurgeons. It guides the appropriate application of LITT, particularly in complex neurosurgical cases in which precision is paramount. Additionally, the histologic outcomes post-LITT provide insights into the long-term effects of therapy, informing both patient prognosis and follow-up care strategies.

### 3.3 Clinical applications and efficacy

LITT’s role in treating primary brain tumors, notably glioblastoma, has been a focal point of numerous studies. In both newly diagnosed and recurrent glioblastoma, LITT has shown potential in extending progression-free survival, offering hope in this challenging area of neuro-oncology (Thomas et al., 2016). The effectiveness of this technique also extends to brain metastases, particularly in cases where stereotactic radiosurgery has failed. Notably, LITT has demonstrated high local control rates for recurrent metastases (Bastos et al., 2020).

Another significant application of LITT is the management of medically refractory epilepsy. The ability of LITT to precisely target epileptogenic foci has resulted in reduced seizure frequency and severity, showcasing its utility across various epileptic pathologies (Johnson et al., 2022). This advancement represents a significant step forward in epilepsy treatment, particularly for patients who do not respond to conventional medical therapies.

The minimally invasive nature of LITT has proven advantageous in accessing lesions situated in challenging locations such as the posterior fossa (Sabahi et al., 2022). Similarly, its application in deep-seated or eloquently located brain areas, which are traditionally associated with high surgical morbidity, marks a significant advancement in neurosurgical techniques. These successes highlight LITT’s role in expanding the boundaries of treatable neurosurgical pathologies.

Although LITT offers safety benefits over open surgery, its complication profile warrants careful consideration. Studies indicate that the complication rates associated with LITT range from 14-35%, encompassing issues such as neurologic complications, wound healing challenges, and psychiatric effects (Viozzi et al., 2021). Specific complications include seizures, infections, hemorrhage, and neurological deficits. Although major morbidity is relatively rare, there is an evident need to refine the understanding of LITT’s toxicity profile of LITT. This will aid in optimizing patient selection and enhancing the overall treatment safety. Continuous monitoring of safety outcomes is crucial as LITT has become more widely adopted in clinical practice.

### 3.4 Role of machine learning

The application of ML techniques in LITT represents a significant stride in neurosurgical innovation. Particularly in preoperative planning, ML algorithms are used to analyze complex patterns in medical imaging and clinical data. This capability allows the development of tailored recommendations that consider unique patient- and lesion-specific characteristics (Li et al., 2019). Such personalized planning is critical for optimizing LITT outcomes.

During the LITT procedure, the ML plays a pivotal role in intraoperative guidance. By processing real-time data, these algorithms can assist surgeons in making informed decisions (Kolasa et al., 2023), thereby enhancing intervention precision. This aspect of ML applications is particularly vital in navigating the intricate anatomy of the brain, where every millimeter counts.

Post-treatment, ML tools extend their utility by tracking patient outcomes (Souza et al., 2023) and modeling the effectiveness of the intervention across diverse patient subgroups. This helps assess the long-term efficacy of LITT and identify potential areas for improvement (Kolasa et al., 2023). Furthermore, the ML-driven analysis of postoperative data can aid in predicting patient recovery trajectories (Souza et al., 2023), enabling more informed follow-up care.

The integration of ML within the LITT framework is more than technological advancement; it represents a fusion of clinical expertise and cutting-edge analytics. As ML continues to evolve, its role in enhancing the clinical adoption of LITT is expected to grow (Viozzi et al., 2021), potentially leading to more effective, efficient, and personalized neurosurgical care.

### 3.5 Surgical planning and trajectory optimization

The incorporation of ML in surgical planning for LITT has marked a significant advancement in neurosurgical precision. ML algorithms are particularly skilled at processing 3D reconstructions and contrast-enhanced images to predict the optimal laser probe trajectories (Li et al., 2019). Precise planning is critical for maximizing the ablation of targets while avoiding eloquent regions that are vital for patient safety (Kolasa et al., 2023). Techniques such as artificial neural networks (ANN) and deep learning models enable accurate recommendations for LITT trajectories and craniotomy planning to meet procedural goals (Li et al., 2019; Souza et al., 2023). These models can be used to analyze anatomical structures and achieve a level of precision that surpasses manual approaches (Kolasa et al., 2023).

Automated ML-based trajectory scoring outperforms the traditional manual planning by neurosurgeons (Kolasa et al., 2019; Kolasa et al., 2023). This shift towards intelligent tools enhances customization for patient-specific anatomy (Souza et al., 2023). Implementing ML facilitates efficient and accurate surgical planning, reducing the time and complexity of LITT preparation (Li et al., 2019; Sabahi et al., 2022). Advancing ML integration can further enhance precision, highlighting the need to refine models using larger datasets (Viozzi et al., 2021; Miranda de Souza et al., 2023). This promises increasingly sophisticated and tailored LITT treatment.

### 3.6 Prediction modeling

ML has proven to be a powerful tool for predictive modeling of LITT outcomes. Advanced techniques, such as regression analysis, decision tree models, and Bayesian networks, have been employed for critical prognosis-related tasks (Kolasa et al., 2023). These models are adept at analyzing complex datasets, allowing for nuanced predictions of LITT outcomes.

A notable application of these ML models is in estimating patient-specific outcomes such as the length of hospital stay post-LITT. These models consider a range of factors, including patient risk profiles and existing comorbidities, to provide individualized predictions (Souza et al., 2023). This level of customization is crucial for preparing both patients and healthcare teams in the postoperative phase.

Furthermore, ML is instrumental in predicting survival. Models incorporating a variety of inputs, from patient demographics and tumor genetics to preoperative neurological status and ablation volumes, have been developed to predict overall and progression-free survival times (Souza et al., 2023). These prognostic models are particularly valuable because they consider the heterogeneity in patient presentations, aiding in distinguishing likely responders from non-responders to LITT therapy.

Although these predictive models show great promise, there remains a need for comprehensive validation studies (Souza et al., 2023). These studies are essential for assessing the accuracy of models across diverse datasets and clinical scenarios. Such validation is a critical step towards ensuring the readiness of these models for real-world clinical applications.

Predictive analytics in LITT offers immense potential for optimizing patient selection and setting realistic expectations for LITT interventions (Viozzi et al., 2021). As these models continue to evolve and undergo validation, they are poised to become an integral part of personalized patient care in neurosurgery, enhancing the decision-making process and improving the overall treatment outcomes.

### 3.7 Evaluation of techniques

As the adoption of LITT has widened across healthcare institutions, ML has become instrumental in evaluating its effectiveness in real-world settings. ML techniques are employed to monitor various aspects of LITT outcomes such as procedure volumes, adherence to established protocols, complication rates, and efficacy benchmarks (Kolasa et al., 2023; Souza et al., 2023). This comprehensive monitoring is essential for understanding how the LITT performs outside controlled study environments.

An innovative approach involves the use of natural language processing methods to gain insights from clinical notes and reports (Kolasa et al. 2023). These methods are adept at extracting valuable information regarding post-LITT metrics such as length of stay, discharge status, and readmission rates. These data provide a deeper understanding of LITT’s impact on patient care and hospital resources. Unsupervised cluster analysis, another key ML technique, has shed light on the variability in patient outcomes following LITT (Miranda de Souza et al., 2023). By identifying different patient profiles, this analysis helps to identify areas where standardized guidelines might be beneficial. This insight is crucial for tailoring LITT to diverse patient needs and improving overall outcomes.

Despite these advancements, challenges remain, particularly regarding the heterogeneity and quality of studies evaluating LITT therapies (Viozzi et al., 2021; Miranda de Souza et al., 2023). Such variability underscores the need for multi-institutional datasets and optimized monitoring methodologies. Addressing these challenges is critical to draw reliable conclusions regarding the efficacy and safety of LITT. To advance the assessment tools for LITT implementation, frameworks that facilitate data acquisition and integration across the entire patient pathway are needed (Miranda de Souza et al., 2023). This holistic approach is essential for evaluating the prognostic capabilities and real-world effectiveness of LITT. Evaluating LITT’s real-world effectiveness of LITT remains a dynamic and evolving area, with ML development at its core (Viozzi et al., 2021). Continued advancements in ML methodologies will be pivotal for enhancing our understanding and application of LITT in clinical settings.

## 4 Discussion

### 4.1 Summary of evidence

This systematic review synthesized studies evaluating applications of LITT as well as integration of ML techniques across various neurosurgical contexts, spanning the treatment of brain tumors, metastatic lesions, radiation necrosis, and more. A review across these studies demonstrates promising efficacy of LITT as a minimally invasive approach, showing survival benefits and progression-free survival extension compared to historical benchmarks across properly selected patients, including those with tumors otherwise unsuitable for resection (Thomas et al., 2016; Bastos et al., 2020).

Analyses also highlight satisfactory safety profiles for LITT relative to open craniotomy access, with complications such as edema, seizures, and infections occurring in 14-35% of procedures but drastically lower rates of serious morbidity compared to operative alternatives (Viozzi et al., 2021). Additionally, emerging data illustrate the potential to enhance multiple aspects of LITT therapy, including surgical planning, predictive modeling, and post-procedural evaluation of real-world outcomes via AI and ML algorithms (Li et al., 2019; Kolasa et al., 2023).

Taken together, LITT exhibits significant utility for ablation of lesions unsuitable for traditional resection, whereas ML tools promise to optimize planning, delivery, and assessment to further improve the precision and personalization of treatment.

However, limitations such as small sample sizes, lack of control groups, heterogeneity in techniques and patient cohorts across studies, and insufficient monitoring methodologies have tempered definitive conclusions regarding standards for best-practice protocols across clinical contexts. Further high-quality comparative effectiveness research through multi-institutional efforts is required to elucidate patient selection criteria, refine procedural guidance, and evaluate incremental benefits over alternative interventions.

### 4.2 Future directions for research

While the findings thus far show promising applications of LITT and ML in neurosurgery, additional high-quality research is vital to clarify outstanding questions, delineate comparative effectiveness, and optimize the integration of these technologies into clinical practice. In particular, large-scale studies leveraging multi-institutional data registries that directly compare survival rates, progression metrics, and complication profiles of LITT to the current gold standard, surgical resection, would greatly help to solidify standardized protocols for appropriate patient selection criteria based on lesion characteristics and comorbidity risk factors.

Furthermore, the integrated analysis of diverse demographic, genomic, imaging, and treatment response data can allow the development of sensitive prognostic models using ML to predict optimal responders to LITT and personalized ablation therapy plans based on individual complication risk. From a technology development perspective, advancing ML algorithms for automated trajectory planning, next-step recommendation systems based on real-time imaging changes, and post-procedural monitoring through seamless EMR integration could accelerate practical clinical adoption. Finally, future work should focus on evaluating emerging combinatorial LITT implementation strategies, such as pairing ablation with adjuvant or neoadjuvant chemotherapy or immunotherapy, which requires systematic investigation regarding synergistic mechanisms and potential efficacy improvements over monotherapies.

In summary, multidisciplinary research that expands retrospective analyses and prospective clinical trials with robust patient cohorts, standardized procedures, ethically sound data governance, and clinical-technological co-innovation represents the next critical leap to firmly establish the role of LITT and ML in transforming routine neurosurgical practice to be less invasive, more quantitatively guided, and maximally personalized for each patient context. Let me know if you need any clarification or have additional suggestions for expanding the overview of impactful future work in this domain.

### 4.3 Implications for clinical practice

The integration of minimally invasive ablation through LITT along with supporting analytics via ML optimizations collectively has salient implications for transforming many facets of traditional neurosurgical practice. Accumulated evidence across this literature suggests expanded personalized treatment options for patients presenting with brain tumors or epileptic lesions in eloquent regions that previously had few alternatives beyond maximal safe resection or biopsy, despite unsatisfactory long-term control. In such complex cases, LITT minimizes morbidity by enabling targeted ablation, thereby avoiding open surgery, which requires more invasive access and risks deficits from damage to adjacent tissues. Shortened hospitalization and evidence of quicker functional recovery further improved the quality metrics across aggregated patient outcomes.

Additionally, ML automation of otherwise time-intensive and experience-dependent planning tasks, such as crafting customized laser probe trajectories, promises greater efficiency in leveraging advances in computational resources to surmount human cognitive constraints. Multi-modal data fusion for predictive analytics at both the population subgroups and individual levels also promotes evidence-based and quantitatively informed neuro-oncology practice over empirical subjectivity alone. Operationalizing such innovations requires building cross-disciplinary coordination between stakeholders, including neurosurgeons, neuroradiologists, ML engineers, clinician data scientists, nursing staff, and technicians. From a healthcare system readiness perspective, prudent LITT adoption further requires upfront and continued investment in both physical infrastructure upgrades, such as intraoperative MRI suites, cloud-based informatics platforms, and clinical workflow redesign with patient-centered data governance.

Balancing implementation costs with quality gains, these transformative technologies portend a future landscape in neurosurgery that promises to be less invasive, more quantitatively guided, efficiently coordinated and thereby maximally personalized for patients based on their unique clinical context.

## 5 Conclusions

This review synthesized the current literature on the applications of LITT and the integration of ML techniques across various neurosurgical indications such as brain tumors, metastases, vascular malformations, and medically refractory epilepsy. Significant retrospective evidence and early prospective studies support the viability of LITT as a minimally invasive treatment option for these pathologies that are not amenable to safe or effective resection, demonstrating satisfactory safety profiles coupled with progression-free and overall survival benefits comparable or favorable to historical benchmarks when applied across properly selected patients. However, small sample sizes, a lack of control groups, variability in LITT protocols, and outcome reporting across studies hinder the highest level of evidence to guide standards and best practices. Upfront investment in multi-center coordinated efforts for high-quality comparative effectiveness trials against surgical resection and stereotactic radiation is critically needed to further consolidate patient stratification criteria and procedural guidance before widespread adoption.

As clinical adoption expands, the emerging integration of ML holds promise in augmenting key facets, such as surgical planning, predictive modeling to set expectations, and monitoring of longitudinal outcomes from LITT applied across wider populations in routine practice. However, the reliability and validity of these tools require rigorous evaluation through nested partnerships between data scientists and clinical domain experts throughout the analytical pipeline. Responsible development and application of explainable AI necessitate shared data standards and governance frameworks that promote transparency, accountability, and ethics to ensure safe and effective translation into clinical practice.

In summary, LITT coupled with ML represents a transformative complementary technology that has immense potential to reshape the management paradigm for some of the most challenging neurosurgical pathologies through minimally invasive and analytics-driven precision approaches. However, this futuristic vision requires a judicious balance of innovation enthusiasm with patient-centric prudence, upheld through sustained appraisal of clinical evidence, outcomes, and experience measures over widespread adoption to forge new frontiers firmly anchored in consistent hippocratic principles.

## Data Availability

All data produced are available through contact with the correspondence email.

---

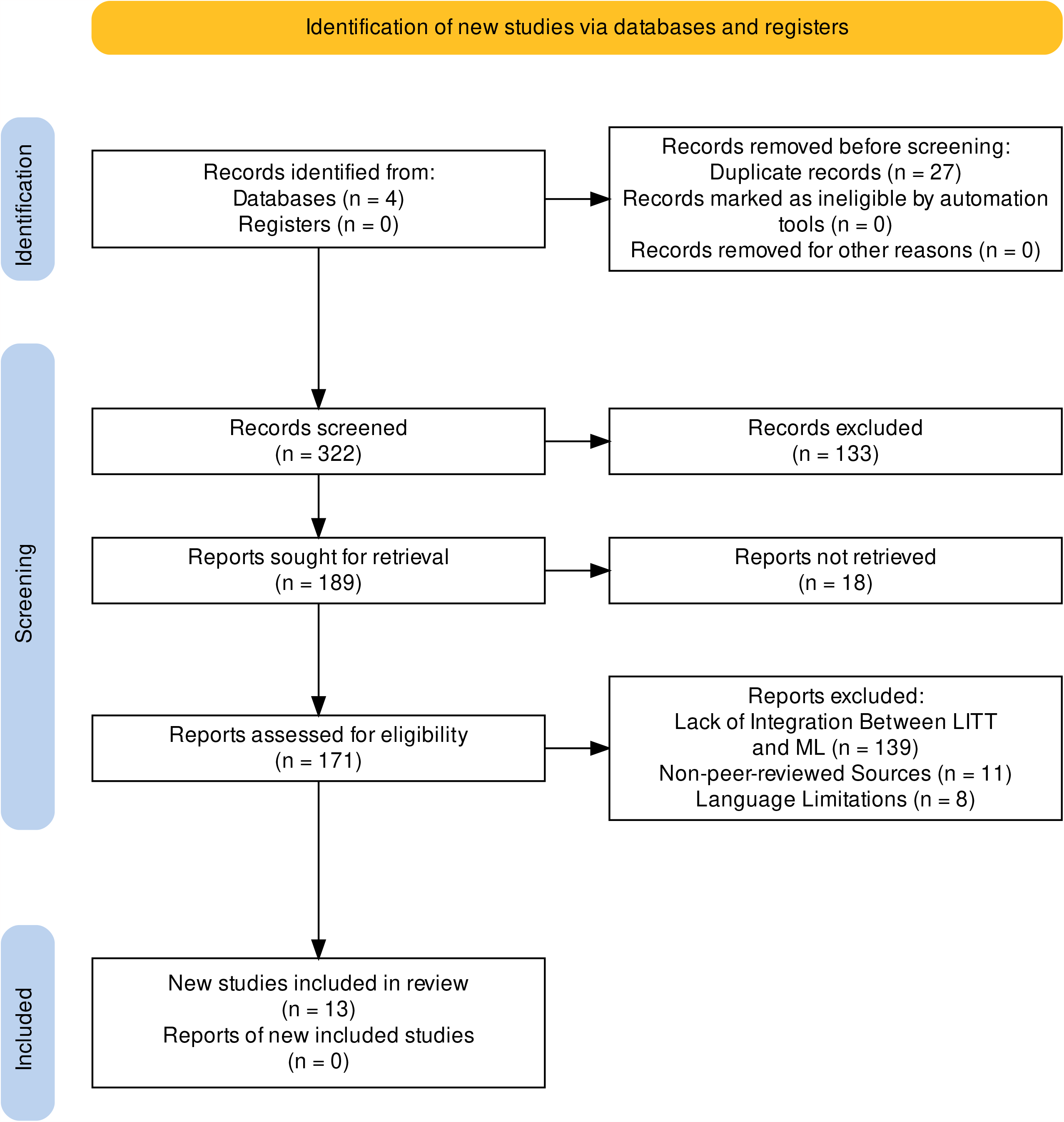

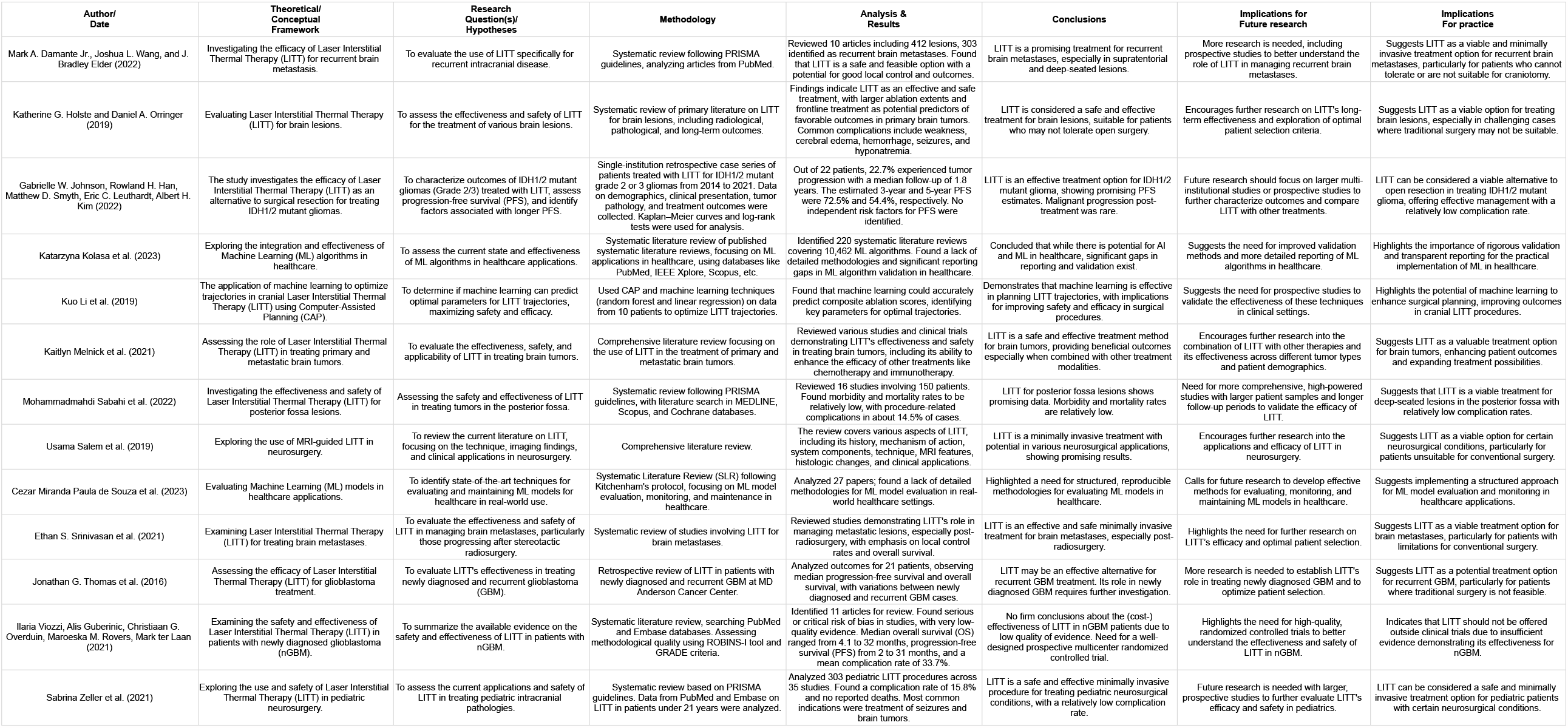

| Author/<br>Date | Theoretical/<br>Conceptual<br>Framework | Research<br>Question(s)/<br>Hypotheses | Methodology | Analysis &<br>Results | Conclusions | Implications for<br>Future research | Implications<br>For practice |
| --- | --- | --- | --- | --- | --- | --- | --- |
| Mark A. Damante Jr., Joshua L. Wang, and J. Bradley Elder (2022) | Investigating the efficacy of Laser Interstitial Thermal Therapy (LITT) for recurrent brain metastasis. | To evaluate the use of LITT specifically for recurrent intracranial disease. | Systematic review following PRISMA guidelines, analyzing articles from PubMed. | Reviewed 10 articles including 412 lesions, 303 identified as recurrent brain metastases. Found that LITT is a safe and feasible option with a potential for good local control and outcomes. | LITT is a promising treatment for recurrent brain metastases, especially in supratentorial and deep-seated lesions. | More research is needed, including prospective studies to better understand the role of LITT in managing recurrent brain metastases. | Suggests LITT as a viable and minimally invasive treatment option for recurrent brain metastases, particularly for patients who cannot tolerate or are not suitable for craniotomy. |
| Katherine G. Holste and Daniel A. Orringer (2019) | Evaluating Laser Interstitial Thermal Therapy (LITT) for brain lesions. | To assess the effectiveness and safety of LITT for the treatment of various brain lesions. | Systematic review of primary literature on LITT for brain lesions, including radiological, pathological, and long-term outcomes. | Findings indicate LITT as an effective and safe treatment, with larger ablation extents and frontline treatment as potential predictors of favorable outcomes in primary brain tumors. Common complications include weakness, cerebral edema, hemorrhage, seizures, and hyponatremia. | LITT is considered a safe and effective treatment for brain lesions, suitable for patients who may not tolerate open surgery. | Encourages further research on LITT's long-term effectiveness and exploration of optimal patient selection criteria. | Suggests LITT as a viable option for treating brain lesions, especially in challenging cases where traditional surgery may not be suitable. |
| Gabrielle W. Johnson, Rowland H. Han, Matthew D. Smyth, Eric C. Leuthardt, Albert H. Kim (2022) | The study investigates the efficacy of Laser Interstitial Thermal Therapy (LITT) as an alternative to surgical resection for treating IDH1/2 mutant gliomas. | To characterize outcomes of IDH1/2 mutant gliomas (Grade 2/3) treated with LITT, assess progression-free survival (PFS), and identify factors associated with longer PFS. | Single-institution retrospective case series of patients treated with LITT for IDH1/2 mutant grade 2 or 3 gliomas from 2014 to 2021. Data on demographics, clinical presentation, tumor pathology, and treatment outcomes were collected. Kaplan–Meier curves and log-rank tests were used for analysis. | Out of 22 patients, 22.7% experienced tumor progression with a median follow-up of 1.8 years. The estimated 3-year and 5-year PFS were 72.5% and 54.4%, respectively. No independent risk factors for PFS were identified. | LITT is an effective treatment option for IDH1/2 mutant glioma, showing promising PFS estimates. Malignant progression post-treatment was rare. | Future research should focus on larger multi-institutional studies or prospective studies to further characterize outcomes and compare LITT with other treatments. | LITT can be considered a viable alternative to open resection in treating IDH1/2 mutant glioma, offering effective management with a relatively low complication rate. |
| Katarzyna Kolasa et al. (2023) | Exploring the integration and effectiveness of Machine Learning (ML) algorithms in healthcare. | To assess the current state and effectiveness of ML algorithms in healthcare applications. | Systematic literature review of published systematic literature reviews, focusing on ML applications in healthcare, using databases like PubMed, IEEE Xplore, Scopus, etc. | Identified 220 systematic literature reviews covering 10,462 ML algorithms. Found a lack of detailed methodologies and significant reporting gaps in ML algorithm validation in healthcare. | Concluded that while there is potential for AI and ML in healthcare, significant gaps in reporting and validation exist. | Suggests the need for improved validation methods and more detailed reporting of ML algorithms in healthcare. | Highlights the importance of rigorous validation and transparent reporting for the practical implementation of ML in healthcare. |
| Kuo Li et al. (2019) | The application of machine learning to optimize trajectories in cranial Laser Interstitial Thermal Therapy (LITT) using Computer-Assisted Planning (CAP). | To determine if machine learning can predict optimal parameters for LITT trajectories, maximizing safety and efficacy. | Used CAP and machine learning techniques (random forest and linear regression) on data from 10 patients to optimize LITT trajectories. | Found that machine learning could accurately predict composite ablation scores, identifying key parameters for optimal trajectories. | Demonstrates that machine learning is effective in planning LITT trajectories, with implications for improving safety and efficacy in surgical procedures. | Suggests the need for prospective studies to validate the effectiveness of these techniques in clinical settings. | Highlights the potential of machine learning to enhance surgical planning, improving outcomes in cranial LITT procedures. |
| Kaitlyn Melnick et al. (2021) | Assessing the role of Laser Interstitial Thermal Therapy (LITT) in treating primary and metastatic brain tumors. | To evaluate the effectiveness, safety, and applicability of LITT in treating brain tumors. | Comprehensive literature review focusing on the use of LITT in the treatment of primary and metastatic brain tumors. | Reviewed various studies and clinical trials demonstrating LITT's effectiveness and safety in treating brain tumors, including its ability to enhance the efficacy of other treatments like chemotherapy and immunotherapy. | LITT is a safe and effective treatment method for brain tumors, providing beneficial outcomes especially when combined with other treatment modalities. | Encourages further research into the combination of LITT with other therapies and its effectiveness across different tumor types and patient demographics. | Suggests LITT as a valuable treatment option for brain tumors, enhancing patient outcomes and expanding treatment possibilities. |
| Mohammadmahdi Sabahi et al. (2022) | Investigating the effectiveness and safety of Laser Interstitial Thermal Therapy (LITT) for posterior fossa lesions. | Assessing the safety and effectiveness of LITT in treating tumors in the posterior fossa. | Systematic review following PRISMA guidelines, with literature search in MEDLINE, Scopus, and Cochrane databases. | Reviewed 16 studies involving 150 patients. Found morbidity and mortality rates to be relatively low, with procedure-related complications in about 14.5% of cases. | LITT for posterior fossa lesions shows promising data. Morbidity and mortality rates are relatively low. | Need for more comprehensive, high-powered studies with larger patient samples and longer follow-up periods to validate the efficacy of LITT. | Suggests that LITT is a viable treatment for deep-seated lesions in the posterior fossa with relatively low complication rates. |
| Usama Salem et al. (2019) | Exploring the use of MRI-guided LITT in neurosurgery. | To review the current literature on LITT, focusing on the technique, imaging findings, and clinical applications in neurosurgery. | Comprehensive literature review. | The review covers various aspects of LITT, including its history, mechanism of action, system components, technique, MRI features, histologic changes, and clinical applications. | LITT is a minimally invasive treatment with potential in various neurosurgical applications, showing promising results. | Encourages further research into the applications and efficacy of LITT in neurosurgery. | Suggests LITT as a viable option for certain neurosurgical conditions, particularly for patients unsuitable for conventional surgery. |
| Cezar Miranda Paula de Souza et al. (2023) | Evaluating Machine Learning (ML) models in healthcare applications. | To identify state-of-the-art techniques for evaluating and maintaining ML models for healthcare in real-world use. | Systematic Literature Review (SLR) following Kitchenham's protocol, focusing on ML model evaluation, monitoring, and maintenance in healthcare. | Analyzed 27 papers; found a lack of detailed methodologies for ML model evaluation in real-world healthcare settings. | Highlighted a need for structured, reproducible methodologies for evaluating ML models in healthcare. | Calls for future research to develop effective methods for evaluating, monitoring, and maintaining ML models in healthcare. | Suggests implementing a structured approach for ML model evaluation and monitoring in healthcare applications. |
| Ethan S. Srinivasan et al. (2021) | Examining Laser Interstitial Thermal Therapy (LITT) for treating brain metastases. | To evaluate the effectiveness and safety of LITT in managing brain metastases, particularly those progressing after stereotactic radiosurgery. | Systematic review of studies involving LITT for brain metastases. | Reviewed studies demonstrating LITT's role in managing metastatic lesions, especially post-radiosurgery, with emphasis on local control rates and overall survival. | LITT is an effective and safe minimally invasive treatment for brain metastases, especially post-radiosurgery. | Highlights the need for further research on LITT's efficacy and optimal patient selection. | Suggests LITT as a viable treatment option for brain metastases, particularly for patients with limitations for conventional surgery. |
| Jonathan G. Thomas et al. (2016) | Assessing the efficacy of Laser Interstitial Thermal Therapy (LITT) for glioblastoma treatment. | To evaluate LITT's effectiveness in treating newly diagnosed and recurrent glioblastoma (GBM). | Retrospective review of LITT in patients with newly diagnosed and recurrent GBM at MD Anderson Cancer Center. | Analyzed outcomes for 21 patients, observing median progression-free survival and overall survival, with variations between newly diagnosed and recurrent GBM cases. | LITT may be an effective alternative for recurrent GBM treatment. Its role in newly diagnosed GBM requires further investigation. | More research is needed to establish LITT's role in treating newly diagnosed GBM and to optimize patient selection. | Suggests LITT as a potential treatment option for recurrent GBM, particularly for patients where traditional surgery is not feasible. |
| Ilaria Viozzi, Alis Guberinic, Christiaan G. Overduin, Maroeska M. Rovers, Mark ter Laan (2021) | Examining the safety and effectiveness of Laser Interstitial Thermal Therapy (LITT) in patients with newly diagnosed glioblastoma (nGBM). | To summarize the available evidence on the safety and effectiveness of LITT in patients with nGBM. | Systematic literature review, searching PubMed and Embase databases. Assessing methodological quality using ROBINS-I tool and GRADE criteria. | Identified 11 articles for review. Found serious or critical risk of bias in studies, with very low-quality evidence. Median overall survival (OS) ranged from 4.1 to 32 months, progression-free survival (PFS) from 2 to 31 months, and a mean complication rate of 33.7%. | No firm conclusions about the (cost-) effectiveness of LITT in nGBM patients due to low quality of evidence. Need for a well-designed prospective multicenter randomized controlled trial. | Highlights the need for high-quality, randomized controlled trials to better understand the effectiveness and safety of LITT in nGBM. | Indicates that LITT should not be offered outside clinical trials due to insufficient evidence demonstrating its effectiveness for nGBM. |
| Sabrina Zeller et al. (2021) | Exploring the use and safety of Laser Interstitial Thermal Therapy (LITT) in pediatric neurosurgery. | To assess the current applications and safety of LITT in treating pediatric intracranial pathologies. | Systematic review based on PRISMA guidelines. Data from PubMed and Embase on LITT in patients under 21 years were analyzed. | Analyzed 303 pediatric LITT procedures across 35 studies. Found a complication rate of 15.8% and no reported deaths. Most common indications were treatment of seizures and brain tumors. | LITT is a safe and effective minimally invasive procedure for treating pediatric neurosurgical conditions, with a relatively low complication rate. | Future research is needed with larger, prospective studies to further evaluate LITT's efficacy and safety in pediatrics. | LITT can be considered a safe and minimally invasive treatment option for pediatric patients with certain neurosurgical conditions. |

